# Robust causal inference for long-term policy decisions: cost effectiveness of interventions for obesity using Mendelian randomization

**DOI:** 10.1101/2020.05.11.20097873

**Authors:** Sean Harrison, Padraig Dixon, Hayley E Jones, Alisha R Davies, Laura D Howe, Neil M Davies

**Affiliations:** MRC Integrative Epidemiology Unit (IEU), Population Health Sciences, Bristol Medical School, University of Bristol, Bristol; Population Health Sciences, Bristol Medical School, University of Bristol, Canynge Hall, 39 Whatley Road, Bristol; Research and Evaluation Division, Public Health Wales NHS Trust, Capital Quarter No.2, Tyndall Street, Cardiff; K.G. Jebsen Center for Genetic Epidemiology, Department of Public Health and Nursing, NTNU, Norwegian University of Science and Technology, Norway

**Keywords:** Obesity, Cost effectiveness, Quality of life years, QALYs, BMI, Body mass index, Mendelian Randomization, UK Biobank, laparoscopic bariatric surgery

## Abstract

**Objectives:** To estimate the cost-effectiveness of interventions to reduce body mass index (BMI) using Mendelian randomization.

**Design:** We estimated the causal effect of differences in BMI on quality-adjusted life years (QALYs) and total healthcare costs using Mendelian randomization and applied our results to policy-relevant questions.

**Setting:** UK Biobank.

**Participants:** 310,913 men and women of white British ancestry aged between 39 and 72 years, followed-up for an average of 8.1 years (6.1 years for secondary care healthcare costs).

**Main outcome measures:** Predicted average QALYs and total healthcare costs per year, and cost-effectiveness of interventions.

**Results:** A unit increase in BMI decreased QALYs by 0.65% of a QALY (95% confidence interval [CI]: 0.49% to 0.81%) per year and increased annual total healthcare costs by £42.23 (95% CI: £32.95 to £51.51) per person. When considering only health conditions usually considered in previous studies (cancer, cardiovascular disease, cerebrovascular disease and type 2 diabetes), we estimated that a unit increase in BMI decreased QALYs by only 0.16% of a QALY (95% CI: 0.10% to 0.22%) per year.

Compared to no intervention and over 20 years, a person in England or Wales aged 40-69 years with a BMI over 35 kg/m^2^ receiving laparoscopic bariatric surgery would have, on average, an increase of 0.92 QALYs (95% CI: 0.66 to 1.17) and a decrease in total healthcare costs of £5,096 (95% CI: £3,459 to £6,852), with a net monetary benefit (at £20,000 per QALY) of £13,936 (95% CI: £8,112 to £20,658).

Restricting volume promotions for high fat, salt and sugar products would, across the 21.7 million adults aged 40 to 69 years in England and Wales, increase QALYs by 20,551 per year (95% CI: 15,335 to 25,301), decrease total healthcare costs by £137 million per year (95% CI: £106 million to £170 million), with a net monetary benefit (at £20,000 per QALY) of £546 million per year (95% CI: £435 million to £671 million).

Between 1993 and 2017 in England and Wales, the increase in BMI of people aged 40 to 69 years led to a decrease of 1.13% of a QALY per person per year (95% CI: 0.90% to 1.38%) and an increase in annual healthcare costs of £69 per person (95% CI: £53 to £84).

Compared to if all people with a BMI above 25 kg/m^2^ aged 40 to 69 years in England and Wales in 2017 had a BMI of 25 kg/m^2^, QALYs are decreased by 580,494 in total per year (95% CI: 457,907 to 717,691) and annual healthcare costs are increased by £3.58 billion (95% CI: £2.75 billion to £4.34 billion).

**Conclusions:** Mendelian randomization can be used to estimate the impact of interventions on quality of life and healthcare costs. The effect of increasing BMI on health-related quality of life is much larger when accounting for 240 chronic health conditions, compared with only a limited selection.

**What is known?:**

- The prevalence of obesity in adults in England and Wales has been increasing over time. Obesity is associated with many chronic illnesses, such as hypertension, coronary artery disease, dyslipidaemia, metabolic liver disease, renal and urological diseases, sleep apnoea, type 2 diabetes, osteoarthritis, psychiatric comorbidity, gastro-oesophageal reflux disease, and some cancers.
- Reliably measuring the impact of obesity on quality of life and healthcare costs is key to informing the cost-effectiveness of interventions that target obesity, helping prioritisation decisions for the allocation of limited resources to address obesity now, as well as how much additional healthcare funding may be required should the trend of increasing obesity continue.
- Previous observational methods of estimating the effect of obesity on quality of life and healthcare costs are subject to bias from confounding and reverse causation; Mendelian randomization is less likely to be affected by these biases.

**What this study adds:**

- We show that Mendelian randomization can be used for policy analysis; in this case, to estimate the effect of an exposure (obesity) on quality of life and healthcare costs. This means we can estimate the cost effectiveness of any specified intervention with potentially less bias than other observational methods.
- This can be especially useful where in the case of obesity, or for other prevalent behaviours with adverse health impacts, such as smoking and alcohol use, where it is difficult, unethical or impossible to randomise participants to the exposure, as well as for interventions where evidence from randomised controlled trials is rare.
- The effect of increasing BMI on health-related quality of life may be larger than previously thought, as existing models may underestimate the effect of BMI on QALYs. This is because we accounted for 240 health conditions, a far larger range than has been previously been considered.

## 1. Introduction

Between 1993 and 2017 in England, the prevalence of obesity in adults aged 40-69 years, defined as a body-mass index (BMI) of over 30 kg/m^2^, rose from 13% to 27% in men and 16% to 30% in women, as estimated by the Health Survey for England (1,2). Obesity is associated with many chronic illnesses, such as hypertension, coronary artery disease, type 2 diabetes, dyslipidaemia, metabolic liver disease, renal and urological diseases, sleep apnoea, osteoarthritis, psychiatric comorbidity, gastro-oesophageal reflux disease, and various cancers (3–7). Reliably measuring the impact on quality of life and the total healthcare cost from obesity is key to informing the cost-effectiveness of interventions that target obesity, and determining how much additional healthcare funding may be required should the trend of increasing obesity continue. For example, prominent recent policy interventions such as the introduction in England of a tax on sugar sweetened beverages were motivated in part by a desire to avoid some of the long-term consequences of obesity on individuals and the health care system (8).

Previous studies examining the cost-effectiveness of interventions for obesity tended to fall into three broad categories: a) randomised controlled trials (RCTs), typically with relatively short-term durations of follow-up (9), b) cohorts, typically retrospective (10–13), and c) decision analytic and related simulation models (10,12,14–18). These studies estimated the impact on quality-adjusted life years (QALYs) and the total healthcare cost of different interventions, such as bariatric surgery, and thus estimated whether the intervention was likely to be cost-effective. **Figure 1, panels a-c** show schematic representations of each type of study, **Box 1** summarises their strengths and limitations, and **Supplementary Information 1** gives more information about each type of study.

Briefly, RCTs with economic evaluations provide causal evidence for cost-effectiveness, but are expensive and time-consuming to perform, while cohort studies are observational and decision analytic simulation models rely on observational evidence that may be subject to confounding and reverse causation that may bias estimates of cost-effectiveness. Decision analytic simulation models also routinely include only a limited selection of health conditions that BMI may affect, meaning the true costs of obesity may be underestimated.

**Figure 1:**
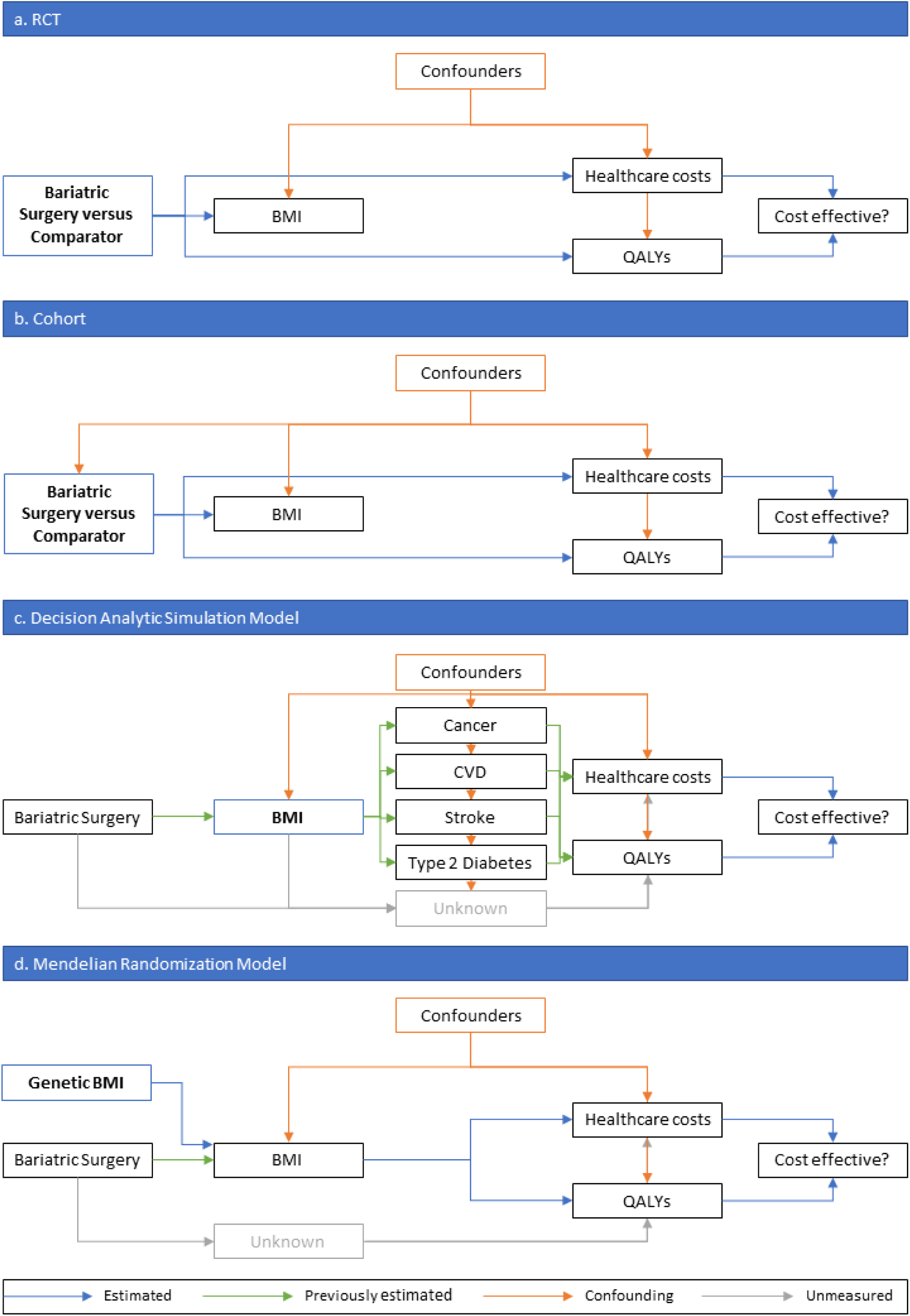
Schematic representation of different methods of estimating cost effectiveness of bariatric surgery. The intervention or exposure for each analysis is in the blue box with bold text. Blue arrows represent what is estimated in each study, while green arrows represent estimates from previous studies used to inform the study. In **panel a**, the estimate of cost-effectiveness is not confounded as the intervention is randomised. In **panel b**, the estimate of cost-effectiveness could be confounded as receiving bariatric surgery is not randomly assigned. In **panel c**, the estimate of cost-effectiveness could be confounded, as could be the estimates from previous studies, there may be effects of bariatric surgery on QALYs and healthcare costs that don’t go through BMI, and there may be effects of BMI on QALYs and healthcare costs that do not go through the modelled health conditions. In **panel d**, the estimate of cost-effectiveness is less likely to be affected by confounding, as genetic variants are randomly distributed within families at conception, though there may be effects of bariatric surgery on QALYs and healthcare costs that don’t go through BMI.

The aim of this study is to elucidate a new approach using Mendelian randomisation (19,20) for estimating the cost-effectiveness of interventions that target BMI, **Figure 1, panel d**. This approach uses observational data, but by using genetic information as an instrumental variable, the risk of bias through confounding and reverse causation is reduced compared with other methods using observational data (21–23). This can give more causal estimates of cost-effectiveness, approximating an RCT of different BMI levels assigned at birth, but with the advantage of estimating at low cost the long-term causal effects of an intervention, rather than shorter-term effects measured during a (usually) limited period of follow-up measured in an economic evaluation conducted alongside an RCT.

In this paper we estimate the causal effect of a unit increase in BMI on both QALYs and total healthcare costs in UK Biobank (24) using Mendelian randomization. We then demonstrate how the results from this approach can be used to estimate the cost-effectiveness of prominent and widely used interventions aimed at reducing BMI (with bariatric surgery and restricting volume promotions for high fat, sugar, and salt products as case studies), estimate the increased healthcare cost of the rise in BMI in England and Wales between 1993 and 2017, and estimate the total cost of the BMI profile of England and Wales in 2017 versus a hypothetical profile where no one has a BMI above 25 kg/m^2^.

#### Box 1

The strengths and limitations of different methods to estimate the cost-effectiveness of interventions

| Method | Strengths | Limitations |
| --- | --- | --- |
| **RCT, with economic evaluation** | • Causal effect estimates | • Expensive<br>• Time-consuming<br>• Often limited follow-up<br>• Study sample may not be representative of target population |
| **Cohort** | • Follow-up may be long<br>• Potentially cheaper than RCTs<br>• A single study can test multiple hypotheses | • Estimates may be biased by confounding and reverse causation (control group not “exchangeable” with intervention group) |
| **Decision analytic simulation models** | • Cheap<br>• Follow-up not limited<br>• Flexible | • Estimates may be biased by confounding and reverse causation<br>• Using only limited health conditions (cancer, cardiovascular disease, cerebrovascular disease and type 2 diabetes) as mediators of the effect of the exposure on the outcome may cause bias<br>• Effect estimates are for a change in the exposure, not an intervention for the exposure, and therefore are best applied to an intervention that affects the exposure across the life course |
| **Mendelian randomization** | • Follow-up may be long<br>• Potentially cheaper than RCTs<br>• A single study can test multiple hypotheses<br>• Estimates less liable to confounding and reverse causation than cohort and decision analytic simulation studies | • Low statistical power; requires very large sample sizes<br>• Effect estimates are for a change in the exposure, not an intervention for the exposure, and therefore are most relevant to proxy an intervention that affects the exposure across the life course |

## 2. Methods

We used Mendelian randomization to estimate the causal effect of BMI on QALYs and total healthcare costs per year. For a guide to Mendelian randomization for clinicians, please see Davies et al. (2018) (20), and for a lay description please see Harrison et al. (2020) (25). Briefly, we generated a polygenic risk score for BMI (a weighted score of genetic risk for higher BMI using common genetic variants), which we used as a proxy for BMI in the Mendelian randomization analyses.

### 2.1 Population

UK Biobank is a population-based health research resource consisting of approximately 500,000 people, who were recruited between the years 2006 and 2010 from 22 centres across the UK (24). Medical data from hospital episode statistics (HES) has been linked to all participants up to 31^st^ March 2017, and primary care (general practice) data has been linked to UK Biobank participants registered with GP surgeries using EMIS Health (EMIS Web) and TPP (SystmOne) software systems, also up to 31^st^ March 2017. The study design, participants and quality control methods have been described in detail previously (26–28). UK Biobank received ethics approval from the Research Ethics Committee (REC reference for UK Biobank is 11/NW/0382). Genotyping information is available in **Supplementary Information 2**, with further information available online (29).

We restricted the main analyses to unrelated individuals of white British ancestry living in England or Wales at recruitment, with a measured BMI value. Full details of inclusion criteria and genotyping are in **Supplementary Information 2**. After exclusions, 310,913 participants remained in the main dataset. Of these, 96,331 (31%) had primary care data covering the full period between recruitment and 31^st^ March 2017 or death, whichever came first.

### 2.2 Polygenic Risk Scores (Instrumental Variables)

We used the Locke 2015 (30) genome-wide association study (GWAS) for body-mass index (BMI) to identify genome-wide significant single nucleotide polymorphisms (SNPs) with strong evidence of association with BMI, defined as having a P value below genome-wide significance (P ≤ 5 × 10^−8^). We clumped the genome-wide significant SNPs at an R^2^ threshold of 0.001 within a 10,000 kilobase window, and proxies were found for all SNPs not in UK Biobank using the European subsample of 1,000 genomes as a reference panel (with a lower R^2^ limit of 0.6) (31). In total, 69 SNPs were used to construct a polygenic risk score (PRS), which we calculated as the weighted sum of the SNP effect alleles for all SNPs associated with BMI, with each SNP weighted by the regression coefficient from the Locke GWAS. **Supplementary Table 1** shows summary data for all SNPs in the PRS. We did not use the more recent 2018 BMI GWAS because this includes the UK Biobank (32), and sample overlap leads to bias towards the observational effect in Mendelian randomization analyses (33).

### 2.3 Exposure and Covariates

We defined BMI as weight in kilograms divided by height in metres squared, and BMI categories using conventional World Health Organization guidelines (34): normal weight as a BMI of between 18.5 kg/m^2^ and 25 kg/m^2^, overweight as a BMI of between 25 kg/m^2^ and 30 kg/m^2^, and obese as a BMI of above 30 kg/m^2^. BMI was estimated at the UK Biobank baseline assessment using measured height and weight.

We used age, sex and UK Biobank recruitment centre reported at the UK Biobank baseline assessment as covariables, as well as 40 genetic principal components derived by UK Biobank to control for population stratification (35).

### 2.4 Estimation of Quality-Adjusted Life Years and Healthcare Costs (Outcomes) Quality-Adjusted Life Years

We predicted health-related quality of life for all participants daily from recruitment to 31 March 2017 using the results from a study by Sullivan et al (36), full details in **Supplementary Information** 3.1. Briefly, we used each of 240 chronic health conditions to predict health-related quality of life for all participants daily from recruitment to 31^st^ March 2017 or death, whichever came first, and averaged over years to estimate quality-adjusted life years (QALYs). **Supplementary Table 2** details all 240 chronic health conditions, including which ICD-9, ICD-10, read v2 and read v3 codes were used for each condition. QALYs are a measure of disease burden, capturing both the quality of life (through preferences over health states, which in this context may be understood as health-related quality of life) and quantity of life (37). A QALY of 1 indicates a full year of perfect health, while a QALY of 0 indicates either a time of no quality of life, or death. QALYs can be negative, implying that death would be preferable to life at a certain time.

Chronic health conditions were recorded in an individual’s primary care data, HES data, or both. As only 31% of participants in this study had primary care data, we used multiple imputation by chained equations to predict both QALYs and primary care healthcare costs (N missing = 214,270, 69%), creating 100 imputed datasets (38). We also imputed Townsend deprivation index (N missing = 342, 0.1%) and whether the participant had ever smoked (N missing =1,064, 0.3%), as these variables were informative but had some missingness. Further details are reported in **Supplementary Information 3.2**.

#### Primary care healthcare costs

We estimated primary care healthcare costs between recruitment and 31 March 2017 from the primary care data as the sum of the cost of prescribed drugs and appointments at a GP practice, see Harrison et al. for more details (39). Briefly, we estimated the cost of prescribed drugs during follow-up using the NHS electronic drug tariff (November 2019 version), adding the cost of each prescription (£1.27 in November 2019) to the cost of each drug (40). In total, we costed 94% of 29,646,535 prescribed drugs, with the remaining drugs either no longer prescribed (and so not costed, n = 392,801, 1.3%) or unmatched to a price (n = 1,392,091, 4.7%). We estimated the cost of each appointment at a GP practice during follow-up at £30, an average of the cost of GP, nurse and other appointments as we could not distinguish between consultation types from the available data (41). We did not consider the cost of diagnostic tests. We divided the total primary care costs by years of follow-up to give the average yearly primary care healthcare costs for each participant.

#### Secondary care healthcare costs

We estimated secondary care healthcare (hospital) costs, in which we converted procedure and diagnosis ICD-10 codes from inpatient episodes into Healthcare Resource Groups, which are assigned a cost (in 2016/17 pounds sterling) for publicly-funded NHS hospitals; see Dixon (2019) for more information (42,43). The data came from HES (for English care providers) and from the Patient Episode Database for Wales (for Welsh providers). Inpatients are those admitted to hospital and who occupy a hospital bed but need not necessarily stay overnight and does not include emergency care or outpatient appointments. We had follow-up data from baseline to 31^st^ March 2015 for secondary care healthcare for all participants in this study. We estimated healthcare costs for those registered in England and Wales only, as the basis for remunerating hospitals in Scotland is different and cannot be combined with data from the other two countries (44).

We estimated the secondary care healthcare cost for each participant between recruitment and 31^st^ March 2015, then divided by the years of follow-up to give the average secondary care healthcare cost per year of follow-up. Secondary care costs were therefore averaged over two fewer years than primary care costs. We increased the value of secondary care healthcare costs by 4.84% to reflect inflation between 2016/17 and November 2019, using data from the NHS cost inflation index, with April to November 2019 inflation estimated at the average annual inflation in the previous 4 years accrued over 8 months (45).

#### Total healthcare costs

We combined the average yearly primary and secondary care healthcare costs for each person to estimate total NHS-based healthcare costs from inpatient hospital care episodes, primary care appointments and primary care drug prescriptions. These costs exclude emergency care, outpatient appointments and private healthcare undertaken in private facilities (private healthcare received in NHS hospitals is included), in addition to diagnostic tests, but still represent a substantial proportion of healthcare costs in England and Wales. Including these other costs would likely increase the size of our effect estimate, but would not alter the direction of the effect.

### 2.5 Main Analysis

We used Mendelian randomization to estimate the causal effect of BMI on QALYs and total healthcare costs per year using the PRS for BMI as an instrumental variable, with age at baseline assessment, sex, UK Biobank recruitment centre and 40 genetic principal components as covariates. We used the ivreg2 package in Stata (version 15.1) with robust standard errors, and tested for weak instrument bias (using F statistics) to assess whether the PRS for BMI was sufficiently associated with measured BMI (46). This Mendelian randomization analysis estimates the mean difference in the outcomes using an additive structural mean model (47-49), interpreted as the average change in each outcome caused by a one kg/m^2^ increase in BMI over all participants. We multiplied the results for QALYs by 100 to give the percentage of a QALY changed per unit increase in BMI.

#### Comparison with multivariable regression approach

We compared the Mendelian randomization estimates with estimates from conventional multivariable linear regression for QALYs and healthcare costs, with age, sex, recruitment centre and 40 genetic principal components as covariates. We performed endogeneity (Hausman) tests (50), in which a low P value indicates there was evidence the Mendelian randomization and multivariable effect estimates were different.

### 2.6 Sensitivity Analyses

**Supplementary Information 3.3** details full methods for all sensitivity analyses.

In brief, we conducted sensitivity analyses to test the Mendelian randomization assumption of no pleiotropy (i.e. that the genetic variants for BMI only affect each outcome through BMI) using summary data for each SNP in the BMI PRS, comprising inverse-variance weighted (IVW), MR Egger (an indicator of directional pleiotropy), weighted median, weighted mode and simple mode analyses (51–53). A low P value in the MR Egger constant would indicate evidence of pleiotropy.

We also re-ran the main analysis stratified by age group (40-49, 50-54, 55-59, 60-64 and 65+ years) and by the World Health Organization BMI categories (normal weight, overweight and obese) (34), to test and account for both non-linearity and a potential interaction between age and BMI in the main effect estimates. We then used non-linear Mendelian randomization to estimate the precise shape of the associations between BMI, QALYs and healthcare costs (54,55). Additionally, we conducted within-family Mendelian randomization to assess whether there was evidence that family structure biased estimates from the main analysis because non-transmitted genetic variants from parents may influence a child’s individual healthcare costs and QALYs in later life (56,57).

We tested whether accounting for prediction uncertainty in QALYs made a material difference to the precision of the main analysis estimates of BMI on QALYs.

Finally, to test whether decision analytic simulation models incorporate enough health conditions to accurately estimate the effect of BMI on QALYs, we estimated whether including only limited health conditions (cancer, cardiovascular disease, cerebrovascular disease and type 2 diabetes) in the prediction of QALYs had a substantial impact on the estimated effect of BMI on QALYs.

### 2.7 Policy Analyses

**Supplementary Information 3.4** details full methods for all policy analyses, and **Supplementary Information 3.5** details a worked example of analysis **d**.

Briefly, we used the results from the Mendelian randomization analyses stratified by age and BMI categories, as well as data and parameter estimates from other studies, to estimate the effect of each of the following on QALYs and healthcare costs for the population aged 40-69 years of England and Wales in 2017 (21.7 million adults):

a. The effect of laparoscopic bariatric surgery in people with a BMI above 35 kg/m^2^
b. The effect of restricting volume promotions for high fat, sugar, and salt (HFSS) foods
c. The effect of the increase in BMI between 1993 and 2017
d. The effect of having the BMI profile of England and Wales in 2017 versus a hypothetical profile where no one has a BMI above 25 kg/m^2^

In example **a** we estimated the net monetary benefit of laparoscopic bariatric surgery as compared to no intervention over 20 years at a cost-effectiveness threshold of £20,000 per QALY and a discount rate for both QALYs and costs of 3.5% per year. We estimated there were 2,741,556 people (12.6%) aged 40 to 69 years with a BMI of 35 kg/m^2^ or above in England and Wales in 2017. We assumed laparoscopic bariatric surgery reduced BMI by 25% (95% CI: 22% to 28%) consistently over 20 years (58,59), and cost £9,549 (60). In example **b** we estimated the net monetary benefit of restricting volume promotions for HFSS foods as compared to no intervention over one year at a cost-effectiveness threshold of £20,000 per QALY. We assumed the intervention reduced Calorific intake by 11-14 Calories per day, that weight is reduced by 0.042 kg per 1 fewer Calorie consumed per day (61,62), and that the intervention had no cost. In example **c** we estimated the change in QALYs and total healthcare costs each year for the change in BMI between 1993 and 2017, and in example **d** we estimated the effect of overweight and obesity on QALYs and total healthcare costs each year. We estimated there were 15,565,145 people (72%) in England and Wales in 2017 with a BMI above 25 kg/m^2^.

We used data from the Health Survey for England in 1993 and 2017 to inform our estimates of the BMI distribution of people in England and Wales (1,2), and data from the Office of National Statistics to inform the age distribution in 2017 (63). We defined the net monetary benefit as the change in QALYs due to the intervention multiplied by a cost-effectiveness threshold (£20,000), minus the change in healthcare costs due to the intervention and the cost of the intervention, including from complications for bariatric surgery for that particular intervention.

### 2.8 Patient and Public Involvement

This study was conducted using UK Biobank. Details of patient and public involvement in the UK Biobank are available online (www.ukbiobank.ac.uk/about-biobank-uk/ and https://www.ukbiobank.ac.uk/wp-content/uploads/2011/07/Summary-EGF-consultation.pdf?phpMyAdmin=trmKQlYdjjnQIgJ%2CfAzikMhEnx6). No patients were specifically involved in setting the research question or the outcome measures, nor were they involved in developing plans for recruitment, design, or implementation of this study. No patients were asked to advise on interpretation or writing up of results. There are no specific plans to disseminate the results of the research to study participants, but the UK Biobank disseminates key findings from projects on its website.

### 2.9 Data and Code Availability

The empirical dataset will be archived with UK Biobank and made available to individuals who obtain the necessary permissions from the study’s data access committees. The code used to clean and analyse the data is available here: https://github.com/sean-harrison-bristol/Robust-causal-inference-for-long-term-policy-decisions.

## 3. Results

In total, we included 310,913 unrelated white British participants from England and Wales in the analysis. These participants had a mean age of 56.9 years (standard deviation (SD) = 8.0 years), mean BMI of 27.4 kg/m^2^ (SD = 4.8 kg/m^2^), a mean follow-up time of 8.1 years (SD = 0.8 years) for primary care healthcare costs and HES data, a mean follow-up time of 6.1 years (SD = 0.8 years) for secondary care healthcare costs, and 10,519 participants died during follow-up (3.4%), see **Table 1**. The median QALY per person per year from the 100 imputed datasets was 0.78 (interquartile range (IQR) = 0.65 to 0.89), compared with 0.97 (IQR = 0.87 to 0.99) based on the HES data alone (nonimputed), reflecting incomplete information on chronic healthcare conditions in HES data. The median total healthcare cost per person per year was £601 (IQR = £212 to £1,217), the median primary care healthcare cost per year was £375 (IQR = £128 to £738), and the median secondary care healthcare cost per year was £88 (IQR = £0 to £494). All cost outcomes were positively skewed.

**Table 1:** Summary demographics UK Biobank

| Variable | All | Men | Women |
| --- | --- | --- | --- |
| N | 310,913 | 144,032 | 166,881 |
| Age at recruitment, years [Mean (SD)] | 56.9 (7.99) | 57.1 (8.10) | 56.7 (7.90) |
| Body Mass Index, kg/m<sup>2</sup> [Mean (SD)] | 27.4 (4.75) | 27.8 (4.22) | 27.0 (5.13) |
| Years of follow-up [Mean (SD)] | 8.1 (0.80) | 8.1 (0.80) | 8.1 (0.80) |
| Participants with complete primary care data [N (%)] | 96,331 (30.98) | 44,671 (31.01) | 51,660 (30.96) |
| Death before 31st March 2017 [N (%)] | 10,519 (3.38) | 6,447 (4.48) | 4,072 (2.44) |
| Qualification: None [N (%)] | 54,874 (17.65) | 25,340 (17.59) | 29,534 (17.70) |
| Qualification: A levels, O level, GCSE or CSE [N (%)] | 122,971 (39.55) | 51,475 (35.74) | 71,496 (42.84) |
| Qualification: NVQ or other [N (%)] | 36,288 (11.67) | 19,873 (13.80) | 16,415 (9.84) |
| Qualification: College or university degree [N (%)] | 96,780 (31.13) | 47,344 (32.87) | 49,436 (29.62) |
| Average QALYs per year (predicted) [Median (IQR)]* | 0.78 (0.65 to 0.89) | 0.78 (0.65 to 0.89) | 0.78 (0.65 to 0.88) |
| Annual Total Healthcare costs [Median (IQR)]* | £601 (£212 to £1,217) | £605 (£206 to £1,240) | £596 (£216 to £1,199) |
| *Results from imputed data, median & IQR are the medians of the 100 imputed medians/IQRs |  |  |  |

### 3.1 Main Analysis

We estimated in the Mendelian randomization that a one kg/m^2^ increase in BMI caused a reduction of 0. 65% of a QALY per year (95% confidence interval [CI]: 0.49% to 0.81%) and a £42.23 increase in total healthcare costs per year (95% CI: £32.95 to £51.51).

#### Comparison with multivariable regression approach

The multivariable adjusted analyses were consistent with the Mendelian randomization analyses, with median P values for endogeneity from imputed datasets 0.31 and 0.52 for QALYs and total healthcare costs respectively, **Table 2**. There was no evidence of weak instrument bias (the F statistic was 5,168). **Figures 2 and 3** show both the Mendelian randomization and multivariable adjusted estimates, for the main analysis, and stratified by sex, BMI category and age category (see **2.6 Sensitivity Analyses**).

**Table 2:** Results from the main Mendelian randomization analysis

| Outcome | Main MR Analysis | Multivariable Adjusted Analysis | P value for Endogeneity |
| --- | --- | --- | --- |
|  | Beta (95% CI) | Beta (95% CI) |  |
| QALYs per year | -0.65% (-0.81% to -0.49%) | -0.71% (-0.73% to -0.69%) | 0.31 |
| Total healthcare costs per year | £42.23 (£32.95 to £51.51) | £39.40 (£38.19 to £40.61) | 0.52 |

**Figure 2:**
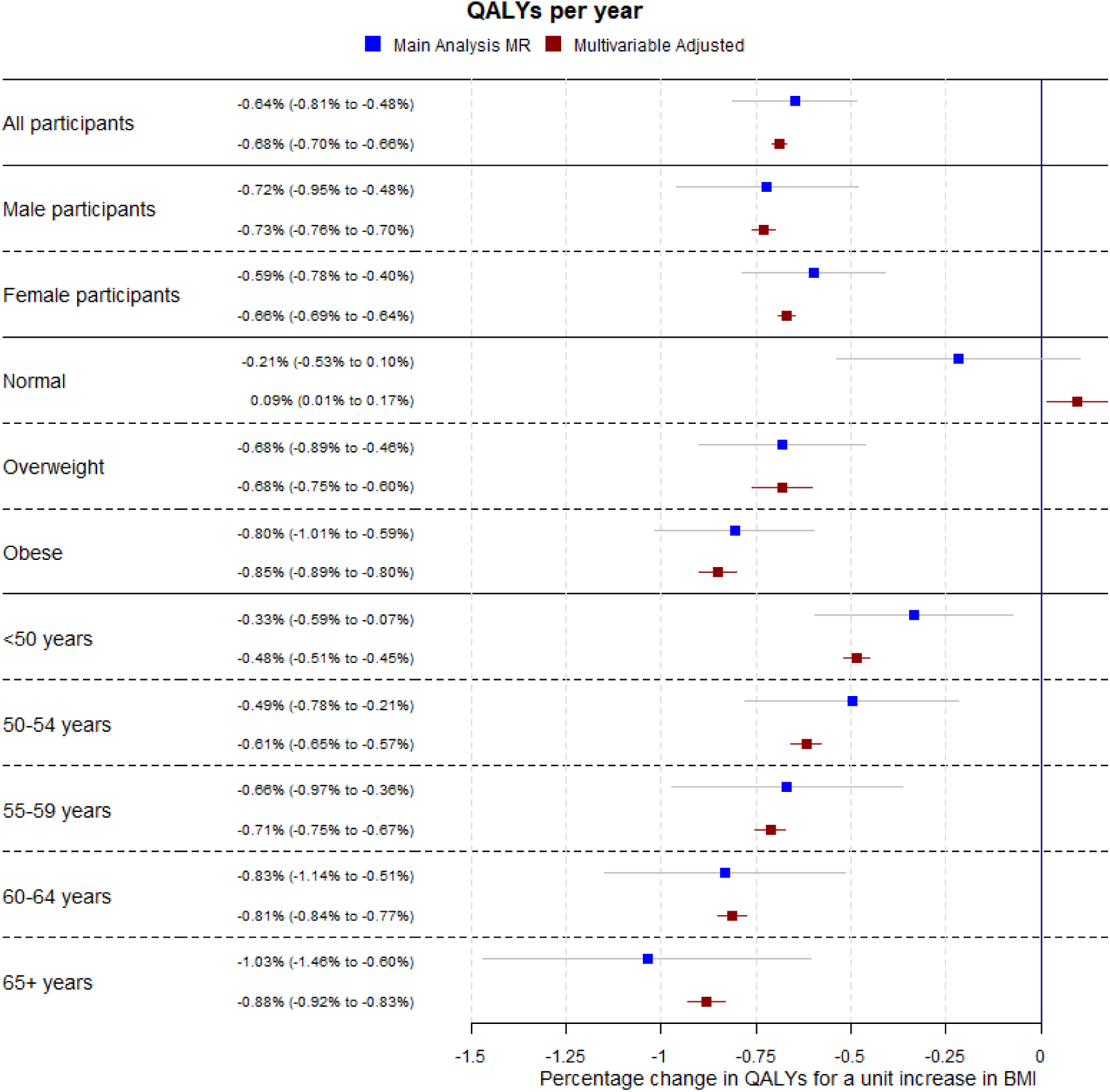
Forest plot showing the estimated effect of a unit increase in BMI on average QALYs per year for the main Mendelian randomization, sex-specific, BMI categorical (where “Normal” is a BMI below 25 kg/m^2^, “Overweight” is a BMI between 25 kg/m^2^ and 30 kg/m^2^, and “Obese” is a BMI of above 30 kg/m^2^) and age categorical analyses.

**Figure 3:**
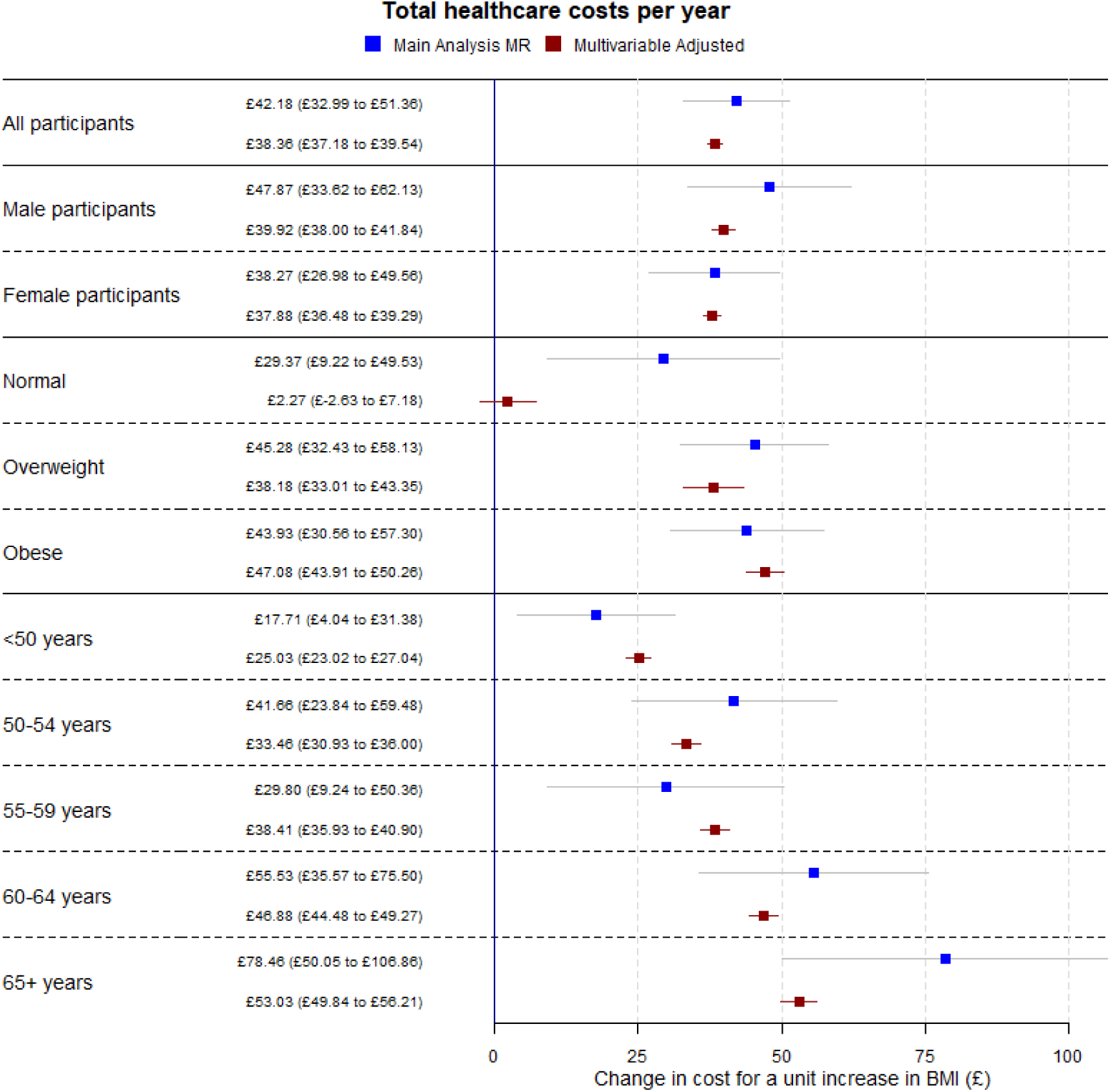
Forest plot showing the estimated effect of a unit increase in BMI on average total healthcare costs per year for the main Mendelian randomization, sex-specific, BMI categorical (where “Normal” is a BMI below 25 kg/m^2^, “Overweight” is a BMI between 25 kg/m^2^ and 30 kg/m^2^, and “Obese” is a BMI of above 30 kg/m^2^) and age categorical analyses.

### 3.2 Sensitivity Analyses

Full results from all sensitivity analyses are in **Supplementary Information 4**.

Briefly, from the summary Mendelian randomization sensitivity analyses, we found little evidence of pleiotropy in the Mendelian randomization estimates, but evidence of heterogeneity in SNP effects using Cochran’s Q value, **Supplementary Table 3**.

We found little difference between the effect estimates when analysing men and women separately; all **Supplementary Tables** have results split by sex. However, we found strong evidence of nonlinearity in the effect of BMI on QALYs, where the effect of the same increase in BMI on QALYs was higher in overweight and obese participants than normal weight participants. There was little evidence of the same non-linearity for total healthcare costs, although this may be due to a lack of power to detect the effects, see **Figures 4 and 5** and **Supplementary Tables 4 and 7**. Additionally, we found evidence for an interaction between BMI and age for both QALYs and total healthcare costs, where the effect of a unit increase in BMI increased as age increased, **Supplementary Table 5**. These results indicate that accounting for sex is not necessary when applying these results to cost-effectiveness analyses but accounting for age and non-linearity of the BMI effect is necessary.

**Figure 4:**
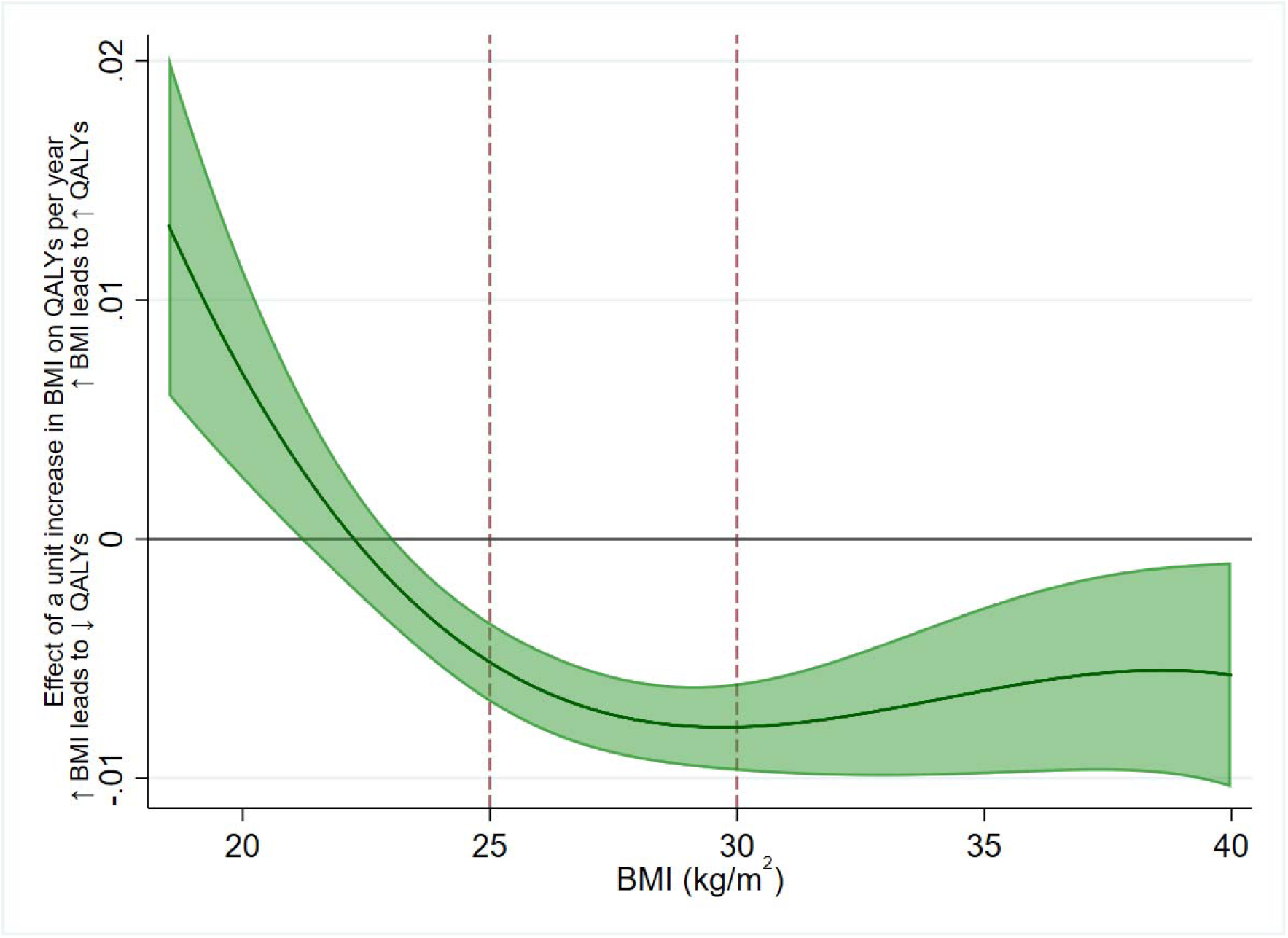
The estimated effect of one kg/m^2^ increase in BMI on QALYs per year, across BMI levels. A positive value indicates an increase in BMI would increase QALYs, and vice versa. An increase in BMI is beneficial to QALYs up to around 22 kg/m^2^, then becomes increasingly detrimental until the effect plateaus in overweight and remains steady relatively in obesity.

**Figure 5:**
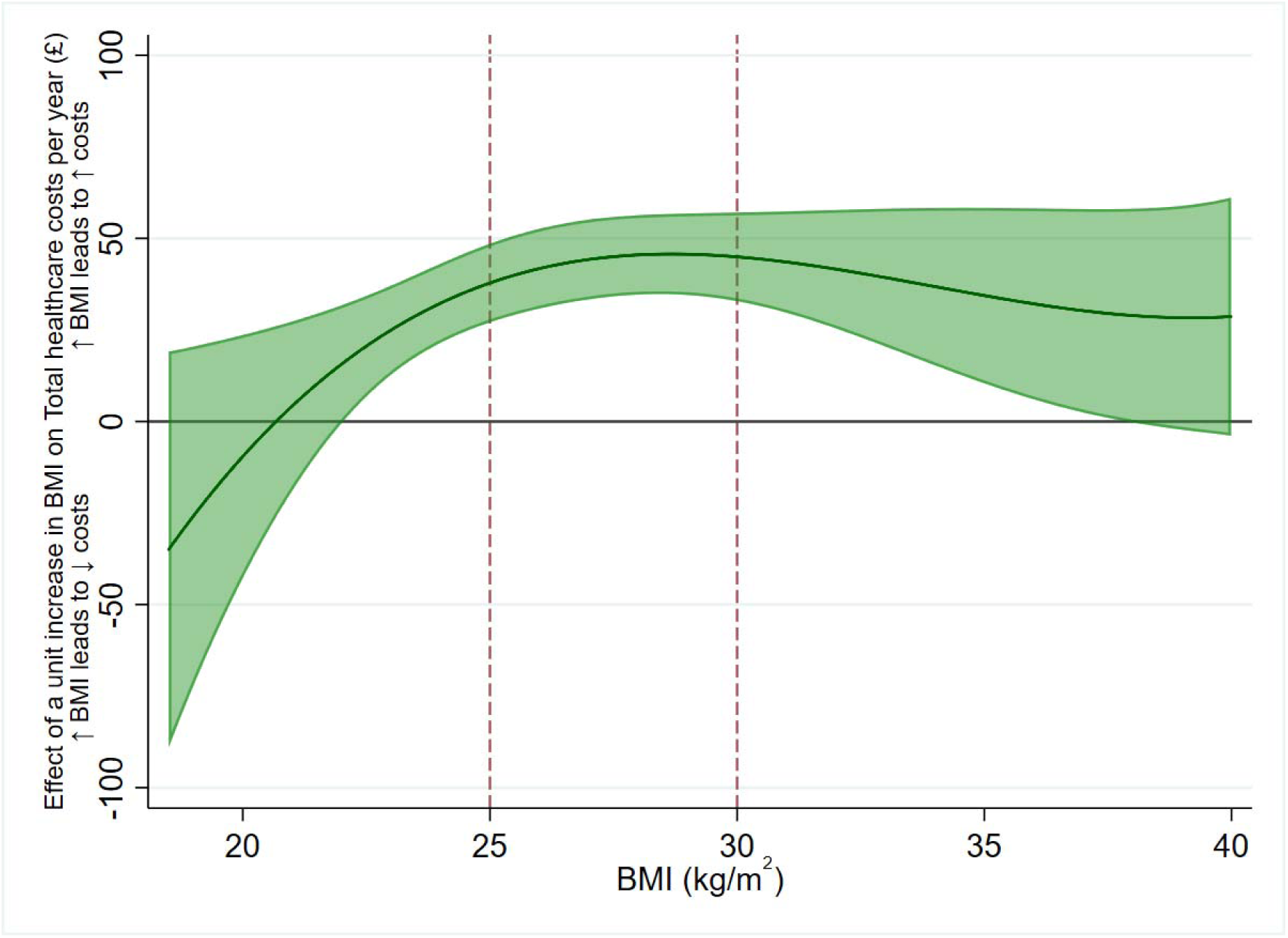
The effect of one kg/m^2^ increase in BMI on total healthcare costs per year, across BMI levels. A positive value indicates an increase in BMI would increase total healthcare costs, and vice versa. Due to the uncertainty in the estimates, there is little statistical evidence of non-linearity in the effect of BMI on total healthcare costs, though descriptively it appears a one kg/m^2^ increase in BMI has a smaller effect on costs in the normal weight category, and a larger effect in overweight and obesity.

The within-family Mendelian randomization analysis estimate for QALYs was very similar to the main analysis estimate but was smaller for total healthcare costs, though both estimates were far less precise, **Supplementary Table 8**. Accounting for the uncertainty in the QALY predictions increased the standard errors of both effect estimates, but not substantially, and did not change the effect estimates, **Supplementary Table 9**.

Predicting QALYs using a limited number of health conditions, as is often done in decision-analytic simulation models drastically reduced the estimated effect of BMI on QALYs, from −0.65% (95%CI: −0.49% to −0.81%) of a QALY to a reduction of 0.16% of a QALY per one kg/m^2^ increase in BMI (95% CI: 0.10% to 0.22%). This indicates BMI affects more health conditions than just cancer, cardiovascular disease, cerebrovascular disease and type 2 diabetes, and these other conditions have a considerable impact on health-related quality of life, **Supplementary Table 10**.

### 3.3 Policy Analyses

#### a. Cost-Effectiveness of an Intervention for BMI

We estimated that 2,741,556 people in England and Wales had a BMI above 35 kg/m^2^ in 2017. Compared to no intervention, over 20 years for each person receiving laparoscopic bariatric surgery we estimated that QALYs would increase by 0.92 (95% CI: 0.66 to 1.17), total healthcare costs would decrease by £5,096 (95% CI: £3,459 to £6,852), and the net monetary benefit (at £20,000 per QALY and £9,549 per intervention) would be £13,936 (95% CI: £8,112 to £20,658). Therefore, laparoscopic bariatric surgery is very likely to be cost-effective over 20 years for people with BMI of 35 kg/m^2^ aged 40 to 69 years in England and Wales. Multivariable adjusted estimates were larger for QALYs and similar for costs, both with greater precision. Full results are in **Supplementary Tables 11 and 12**.

#### b. Cost-Effectiveness of restricting volume promotions for high fat, sugar, and salt (HFSS) products

We estimated that restricting volume promotions for HFSS products would increase QALYs by 20,551 per year (95% CI: 15,335 to 25,301), decrease total healthcare costs by £137 million per year (95% CI: £106 million to £170 million), and would have a net monetary benefit (at £20,000 per QALY and no intervention cost) of £546 million per year (95% CI: £435 million to £671 million). The intervention would therefore almost certainly be cost effective, relative to doing nothing. Multivariable adjusted estimates were larger for QALYs and similar for costs, both with greater precision. Full results are in **Supplementary Tables 13 and 14**.

#### c. Estimation of the Effect of the Population Change in BMI Between 1993 and 2017

Mean BMI increased from 26.7 kg/m^2^ to 28.6 kg/m^2^ between 1993 and 2017 in people aged between 40 and 69 years in England and Wales. The rise in BMI was more pronounced in people with obesity than people with a normal weight, see **Supplementary Table 15**.

We estimated that between 1993 and 2017, the increase in BMI led to an average decrease in QALYs of 1.13% per person per year (95% CI: 0.90% to 1.38%), or a decrease of 246,390 QALYs in total per year (95% CI: 196,231 to 300,481) and an increase in total healthcare costs of £69 per person per year (95% CI: £53 to £84), or £1.50 billion in total per year (95% CI: £1.15 billion to £1.82 billion), giving a combined cost (at £20,000 per QALY) of £312 per person per year (95% CI: £235 to £347), or £6.39 billion (95% CI: £5.12 billion to £7.54 billion). This indicates an intervention which could reduce the BMI of the population of England and Wales to 1993 levels would likely be cost effective if it cost less than £5.12 billion per year. Multivariable adjusted estimates were larger for QALYs and similar for costs, both with greater precision. Full results are in **Supplementary Tables 16 and 17**.

#### d. The Cost of Being Overweight and Obese in 2017

We estimated that, compared to if all people with a BMI above 25 kg/m^2^ aged 40 to 69 years in England and Wales in 2017 had a BMI of 25 kg/m^2^, QALYs are decreased by 3.73% per person (with a BMI above 25 kg/m^2^) per year (95% CI: 2.94% to 4.61%), or a decrease of 580,494 QALYs in total per year (95% CI: 457,907 to 717,691), and an increase in total healthcare costs of £230 per person per year (95% CI: £176 to £279), or £3.58 billion in total per year (95% CI: £2.75 billion to £4.34 billion), giving a combined cost (at £20,000 per QALY) of £973 per person per year (95% CI: £773 to £1160), or £15.1 billion (95% CI: £12.0 billion to £18.1 billion). Multivariable adjusted estimates were larger for QALYs and similar for costs, both with greater precision. Full results are in **Supplementary Tables 18 and 19**.

## 4. Discussion

In this study, we have shown that cost effectiveness of clinical and policy interventions can be estimated using Mendelian randomization. We estimated the effect of a unit increase in BMI on average QALYs and total healthcare costs per year in UK Biobank, which showed that increasing BMI is detrimental to both QALYs and healthcare costs in people with overweight and obesity. The effect estimates were relatively stable once BMI had reached 25 kg/m^2^, implying that the detrimental effect of an increase in BMI is very similar whether a person has overweight or obesity, and, conversely, a reduction in BMI is similarly beneficial. We used these estimates to show that bariatric surgery and the restriction of volume promotions for HFSS products are likely cost-effective relative to a “no intervention” comparator and estimated the costs of the increase to BMI over time and having overweight and obesity in 2017. We emphasise that all cost-effectiveness estimates are relative to a “no intervention” comparator.

We have demonstrated how Mendelian randomization can be useful for estimating the impact on quality of life and healthcare costs of either an exposure or intervention that is difficult, unethical or impossible to randomise (e.g. smoking, alcohol intake), or for interventions where long-term cost-effectiveness evidence from RCTs is rare or not generalisable (e.g. bariatric surgery). While in this study the conventional multivariable adjusted estimates not using genetic information were mostly similar to the Mendelian randomization estimates, this could be due to larger uncertainty in the Mendelian randomization estimates, and there is no guarantee that other exposures will be similar.

### Strengths and limitations

The estimates of the effect of BMI on QALYs and costs from Mendelian randomization are likely less biased by confounding and reverse causation than either cohort studies or decision analytic simulation models using observational effect estimates (20). UK Biobank has many participants with comprehensive information about costs and disease states over many years. While the corresponding conventional multivariable adjusted estimates were generally consistent with the Mendelian randomization estimates for all outcomes, the Mendelian randomization estimates showed some detrimental effect of increasing BMI even in participants with BMI close to the top end of the normal weight category, while the conventional estimates did not, which could reflect bias in the conventional estimates.

This method of estimating the effect of a risk factor on QALYs and costs can be extended to other risk factors with causal genetic components, and also provide evidence for the causal effects of health conditions on healthcare costs and QALYs. This may be useful for health conditions that are strongly influenced by risk factors that affect other health conditions where the effect of the condition would otherwise be confounded by the risk factor, such as cardiovascular disease.

However, Mendelian randomization relies on assumptions that cannot be proven (20), as is the case with all types of instrumental variable analysis and other forms of observational policy evaluation. There was evidence for heterogeneity between SNPs for all outcomes, though in general the summary Mendelian randomization sensitivity estimates were consistent with the main estimates, and there was little evidence of directional pleiotropy from the MR Egger regression. As the outcomes were not biological, the exclusion restriction assumption (i.e. that any genetic variant affects the outcome only through the exposure) may not hold for all the genetic variants (i.e. that the genetic variant affects the outcome only through the exposure).

These estimates represent a lifetime exposure to a genetic influence on BMI, and thus cannot be interpreted directly as the expected effect of an intervention at a specific age. In general, as the age at which a person received an intervention increases, the effect estimates would likely reduce. This is because the mechanisms by which BMI affects health may be cumulative over time, and so even if BMI were lowered in older age, some residual detrimental effect of previously high BMI may remain.

It is therefore likely that our estimates of the impact of BMI on costs and QALYs are best applied to population level interventions that aim to reduce BMI across all age groups. This limitation is also present in decision analytic simulation models of cost-effectiveness, though not RCTs or cohort studies. Our estimates may also underestimate the true effect as people in England and Wales now may have had larger BMI values earlier in life than previously, increasing the length of exposure to obesity.

For all policy examples, we require the stable unit treatment value assumption for causal inference, this assumption requires that genetic change in BMI is equivalent to a change in BMI by other means, e. g. by bariatric surgery or reducing Caloric intake of high fat, sugar and salt foods. This assumption is not testable. Mendelian randomization analyses can also be interpreted as estimates of a “local average treatment effect”, by assuming that changes in the genetic variants affecting BMI affect all participants in UK Biobank in the same direction (monotonicity). This is assumption also cannot be tested, and deviations from monotonicity could bias effect estimates.

The analyses accounting for QALY prediction error were consistent with the main analysis, although less precise. We also had to impute primary care costs and QALYs as only a limited section of UK Biobank had primary care data, which limited statistical power but was unlikely to have biased the results, rather, the complete case analysis would likely have been biased results, since the distribution of GP software systems allowing linkage of primary care data is unlikely to be random.

The healthcare costs were estimated from observed hospital episodes, drug prescriptions and appointments from primary care. Follow-up was two years shorter for secondary care costs than primary care costs, but as we averaged the costs this should not have materially affected the results. Additionally, we did not capture all healthcare costs as we did not have access to private healthcare costs not incurred in NHS settings, or data for emergency care or outpatient appointments (which are not linked to the UK Biobank cohort), and did not consider the cost of diagnostic tests in primary care, likely therefore under-estimating the total cost of increasing BMI. In contrast, participants in UK Biobank may have different access to healthcare than the country on average, which may have biased our estimates of the effect of BMI on costs. Finally, BMI may have interacted with the use of both state and private healthcare, potentially biasing the results in either direction.

Despite its size, UK Biobank is not representative of the UK population as participants tend to be wealthier and healthier compared to the country on average (64). It therefore likely that we have underestimated the true costs of BMI, as wealthier and healthier people may be more resistant to any detrimental effects of increased BMI. As obesity is more common in lower socioeconomic groups (65), our results indicate that obesity is likely a cause of inequalities in quality of life.

Although Mendelian randomization is likely to be less affected by confounding and reverse causality than conventional multivariable adjusted analyses, an important potential source of bias in these analyses is family-level effects. Recent evidence suggests that assortative mating and dynastic effects can lead to bias in Mendelian randomization effect estimates (56), though within-family Mendelian randomization studies can account for some of these biases. Our within-family sensitivity analyses showed that the effect of BMI on QALYs was consistent with the main analysis, though the effect of BMI on total healthcare costs was reduced. However, statistical power was limited in these analyses, and confidence intervals were wide. Additionally, there is evidence of a geographic structure in the UK Biobank genotype data that cannot be accounted for using adjustment for principal components, which may also have biased our analyses (66).

## 5. Conclusion

Mendelian randomization can be used to estimate the effect of an exposure on quality of life and healthcare costs. We used this approach to estimate the cost effectiveness of interventions aimed at reducing BMI. This approach could be especially useful where it is difficult, unethical or impossible to randomise participants to an exposure such as obesity or for prevalent behaviours with adverse health impacts such as smoking or alcohol use, or where RCT evidence is rare for an intervention. Results from such studies are likely of benefit to both policy and the NHS. In future studies, we will use this method to assess the costs of different risk factors for poor health.

The effect of increasing BMI on health-related quality of life may be larger than previously thought, as decision analytic simulation models may underestimate the effect of BMI on QALYs by using only limited health conditions are intermediates.

## Acknowledgements

This research has been conducted using the UK Biobank Resource under Application Number 29294. Quality Control filtering of the UK Biobank data was conducted by R. Mitchell, G.Hemani, T.Dudding, L.Paternoster as described in the published protocol (doi:10.5523/bris.3074krb6t2frj29yh2b03x3wxj). The MRC IEU UK Biobank GWAS pipeline was developed by B.Elsworth, R.Mitchell, C.Raistrick, L.Paternoster, G.Hemani, T.Gaunt (doi: 10.5523/bris.pnoat8cxo0u52p6ynfaekeigi). The Medical Research Council (MRC) and the University of Bristol support the MRC Integrative Epidemiology Unit [MC_UU_12013/1, MC_UU_12013/9, MC_UU_00011/1]. NMD is supported by an Economics and Social Research Council (ESRC) Future Research Leaders grant [ES/N000757/1] and the Norwegian Research Council Grant number 295989. LDH is supported by a Career Development Award from the UK Medical Research Council (MR/M020894/1). PD acknowledges support from a Medical Research Council Skills Development Fellowship (MR/P014259/1).This work is part of a project entitled ‘social and economic consequences of health: causal inference methods and longitudinal, intergenerational data’, which is part of the Health Foundation’s Social and Economic Value of Health Programme (Grant ID: 807293). The Health Foundation is an independent charity committed to bringing about better health and health care for people in the UK. No funding body has influenced data collection, analysis or its interpretation. This publication is the work of the authors, who serve as the guarantors for the contents of this paper.

## Author contributions

**Sean Harrison**: Conceptualization, Methodology, Software, Formal analysis, Data curation, Writing – original draft, review & editing, Visualization

**Padraig Dixon**: Conceptualization, Methodology, Software, Writing – review & editing

**Hayley E Jones**: Conceptualization, Methodology, Writing – review & editing, Supervision, Funding acquisition

**Alisha R Davies**: Writing – review & editing, Funding acquisition

**Laura D Howe**: Conceptualization, Methodology, Writing – review & editing, Funding acquisition

**Neil M Davies**: Conceptualization, Methodology, Writing – review & editing, Supervision, Funding acquisition

## Transparency Statement

Transparency statement: The lead author (the manuscript’s guarantor) affirms that this manuscript is an honest, accurate, and transparent account of the study being reported; that no important aspects of the study have been omitted; and that any discrepancies from the study as planned (and, if relevant, registered) have been explained.

